# The Carrier Delivery-Assist Catheter in Stroke Thrombectomy

**DOI:** 10.64898/2026.04.27.26351898

**Authors:** Jaydevsinh Dolia, Theja Yelam, Jonathan Grossberg, Savio Batista dos Reis, Aqueel Pabaney, Mithilesh Siddu, Daniel Vela-Duarte, Brian Jankowitz, Omar Tanweer, Jordan Xu, Hugo Cuellar-Saenz, Rahul Shah, Isaac Abecassis, Dale Ding, Tapan Mehta, Sunil Sheth, Joseph Samaha, Sami Al Kasab, Kevin Shah, Michael Froehler, Amaan Ali, Ameer Hassan, Samantha Miller, Jeffrey Miller, Tareq Kass-Hout, Rami Z. Morsi, Kaustubh Limaye, Pedro Martins, Diogo Haussen

## Abstract

**Introduction:** Delivering large-bore aspiration catheters through tortuous anatomy remains challenging during mechanical thrombectomy (MT). The Carrier delivery-assist catheter (DAC) was designed to facilitate aspiration catheter navigation, but multicenter data remain limited. We evaluated the efficiency and safety of the Carrier DAC.

**Methods:** We performed a multicenter retrospective study of prospectively collected data from patients undergoing MT at 15 U.S. Comprehensive Stroke Centers (September 2024–September 2025). Co-primary endpoints were puncture-to-clot engagement time and first-pass effect (FPE; eTICI 2c–3). A pre-specified single-center analysis compared upfront contact aspiration using the Carrier DAC versus standard 0.021″ microcatheter techniques with identical aspiration catheter sizes.

**Results:** The multicenter cohort included 211 Carrier-assisted MTs. Median aspiration catheter inner diameter was 0.071″, with super-bore catheters used in 5.7%. Median puncture-to-clot time was 12 minutes, and FPE was achieved in 50.7%. Median puncture-to-reperfusion time was 20 minutes, and mFPE occurred in 74.4%. Parenchymal hematoma and subarachnoid hemorrhage occurred in 11.8% and 6.6%, respectively. Cavernous tortuosity did not affect primary endpoints. The single-center analysis included 242 patients. Carrier use was associated with shorter puncture-to-clot times and numerically higher FPE rates without increased hemorrhagic complications.

**Conclusions:** The Carrier DAC enables efficient navigation of large-bore aspiration catheters and may reduce procedural time while maintaining procedural safety. Prospective studies are warranted.

## INTRODUCTION

Mechanical thrombectomy (MT) has become the standard of care for large vessel occlusion (LVO) acute ischemic stroke (AIS), as established by pivotal randomized trials primarily utilizing stent-retrievers.^1-2^ Subsequently, contact aspiration (CA) has been shown to offer comparable performance, leading to a significant increase in its adoption in recent years. ^3-5^ Technological advancements in catheter design have enabled the development of aspiration catheters with larger inner diameters (ID), which have been associated with improved rates of first-pass effect (FPE).^6-9^ However, consistent delivery large-bore and super large bore catheters through tortuous anatomy remains challenging. Adjunctive devices such as macrowires and microwires/microcatheters may not allow for successful navigation, and the addition of smaller sized aspiration catheters may increase procedural cost.^10-12^

The delivery-assist catheters (DACs) have emerged as a class of devices intended to facilitate large and super-large bore catheter navigation to the clot.^13^ DACs are hydrophillically coated, feature distal tapering, and have a slightly smaller outer diameter relative to the primary aspiration catheter. They function as a coaxial support system providing a stable conduit, serving as a rail for more consistent navigation and improved deliverability of large and super large bore aspiration catheters.^14^ The Carrier DAC (Balt, Irvine, CA) is specifically engineered with a low-profile design and tapered distal tip, aiming to minimize the “ledge effect” commonly encountered in tortuous anatomy/branching points and to reduce the navigation-related risk of vessel injury. The Carrier features a 0.021-inch inner diameter, enabling compatibility with 0.014- and 0.018-inch guidewires, and incorporates radiopaque markers for reliable positioning. It is designed to integrate with a wide range of aspiration catheters, allowing operators to maintain flexibility in device selection and thrombectomy strategy.^15^

In this multicenter retrospective study, we evaluate the initial procedural performance and safety of the Carrier DAC in MT. Additionally, a single center comparative analysis explores how the Carrier DAC performs relative to conventional microcatheter-microwire assisted delivery.

## METHODS

### Multicenter Study Design and Population

We conducted a multicenter, retrospective study of a prospectively collected data on patients with LVO treated with MT at 15 Comprehensive Stroke Centers (CSC) in the United States spanning September 2024 to September 2025. The study was approved by the local institutional review boards at each participating site, with a waiver of informed consent due to the retrospective design.

### Inclusion Criteria and Device Use

Inclusion criteria consisted of consecutive patients with anterior or posterior circulation occlusions who underwent MT using the Carrier DAC for the first-pass attempt. Use of the Carrier DAC was based on individual operator preference, including technique selection regarding microwire use versus direct advancement without a microwire. Small (outer diameter - OD 0.059”), medium (OD 0.062”) or large (OD 0.069”) Carrier DACs sizes were utilized. Cases where the Carrier DAC facilitated stent retriever in addition to aspiration were also included.

### Endpoints

The co-primary endpoints were time-to-clot (time window between arterial puncture and clot engagement) and first pass effect (FPE - eTICI2c-3 with the first device pass). Secondary endpoints included time from puncture-to-successful reperfusion (defined as mTICI2b-3), rates of parenchymal hematoma (PH1 and PH2 as per ECASSIII criteria), ^16^ and modified first pass effect (mTICI2b-3 with the first pass).

### Single Center Comparative Analysis: Carrier DAC vs Standard Microcatheter

A pre-specified single-center comparative analysis of patients with LVO (intracranial internal carotid artery-ICA, middle cerebral artery-MCA M1 or M2 and basilar artery) treated with primary CA technique spanning January 2023 to January 2025 was pursued. The cohort of contact aspiration via Carrier DAC was compared to a population with identical aspiration catheter devices (brand and model, including 0.070”, 0.071”, 0.074”, or 0.088” ID sizing) navigated via standard technique using 0.021” microcatheter and 0.014-18” microwire. The same endpoints utilized for the main cohort were used.

### Statistical Analysis

Demographic and clinical data were reported as median [IQR] and categorical variables are reported as proportions. Between group comparisons were made with Student t-test/Mann-Whitney U, Chi-square/Fisher exact test. Significance was set at P<0.05. Statistical analyses were conducted using R Studio software (version 4.4.1, R Foundation for Statistical computing, Vienna, Austria).

For the multicenter cohort, descriptive statistics were used. Adjusted linear and logistic regression mixed models evaluating the association between centers for puncture-to-clot and FPE, respectively, were pursued (centers as random effects and adjusted for age, catheter size, occlusion site, and vascular access) and the intraclass correlation coefficient (ICC) was calculated as the proportion of total variance attributable to centers. A sensitivity analysis evaluating Type 1–2 vs Type 3–4 cavernous ICA tortuosity^17^ was pursued to determine the performance of Carrier in complex vascular anatomy. Adjusted linear and logistical regression models evaluating the association of type of cavernous carotid tortuosity with the primary and secondary endpoints were pursued (adjusted for age, NIHSS, catheter size, and occlusion site) were pursued.

Adjusted linear and logistical regression models comparing Carrier DAC vs standard microcatheter technique for the primary and secondary endpoints were pursued (adjusted for age, catheter size, and occlusion site).

## RESULTS

### Multicenter Study Population

The multicenter experience included 211 stroke Carrier-mediated stroke thrombectomies. The baseline characteristics represent a typical thrombectomy cohort (**Table 1**). The median age was 70 (IQR, 69-79) years, median admission NIHSS 16 (11-21) and ASPECTS 8 (IQR,7-10). Occlusion site included intracranial ICA in 18.9% of cases, MCA-M1 64%, MCA-M2 10%, MCA-M3 1.4%, anterior cerebral artery in 0.9%, posterior cerebral artery in 0.5% and basilar artery 4.3% with 5.2% representing tandem occlusions. Two-thirds of MT cases utilized general anesthesia and 89.6% transfemoral access. The median aspiration catheter ID was 0.071” (IQR, 0.071-0.072”) and 5.7% of MTs utilized super-bore (ID≥ 0.084”). Carrier DAC-facilitated stent retriever delivery followed by contact aspiration (combined technique) was used in 10% of cases.

**Table 1:** Baseline Characteristics.

| **Full Carrier Cohort** | **n = 211** |
| --- | --- |
| **Age [years]** | 70 [62-79] |
| **Ethnicity** |  |
| Black | 72 [34.1%] |
| White | 100 [47.4%] |
| Hispanic | 27 [12.8%] |
| Asian | 8 [3.8%] |
| Other | 3 [1.4%] |
| **Hypertension** | 161 [76.3%] |
| **Dyslipidemia** | 120 [56.9%] |
| **Diabetes** | 73 [37%] |
| **Atrial Fibrillation** | 76 [36%] |
| **Smoking [Current]** | 43 [20.4%] |
| **NIHSS** | 16 [11-21] |
| **ASPECTS** | 8 [7-10] |
| **IV Thrombolysis** | 54 [25.6%] |
| **Cavernous ICA tortuosity** |  |
| Type 1 | 90 [42.7%] |
| Type 2 | 53 [25.1%] |
| Type 3 | 36 [17%] |
| Type 4 | 23 [10.9%] |
| **Extracranial ICA tortuosity** |  |
| Tortuous | 88 [41.7%] |
| Coiled | 18 [8.5%] |
| Straight | 62 [29.4%] |
| Kinked | 34 [16.1%] |
| **Occlusion Site** |  |
| Internal carotid artery | 38 [18%] |
| MCA M1 | 127 [60.2%] |
| MCA M2 | 20 [9.5%] |
| MCA M3 | 3 [1.4%] |
| Basilar | 9 [4.3%] |
| Anterior Cerebral Artery | 2 [0.9%] |
| Posterior Cerebral Artery | 1 [0.5%] |
| Tandem | 11 [5.2%] |

Legend: NIHSS - National Institutes of Health Stroke Scale, ASPECTS – Alberta Stroke Program Early CT Score, ICA – Internal Carotid Artery, MCA- Middle Cerebral Artery.

### Procedural and Efficacy Outcomes

The analysis of co-primary endpoints showed a median time from puncture-to-clot of 12 (IQR,9-19) minutes and FPE (eTICI2c-3) rate of 50.7%(**Table 2**). In terms of secondary endpoints, the median time from puncture to successful reperfusion was 20 (IQR,13-32) minutes and the mFPE rate 74.4%. The influence of centers in the observed puncture-to-clot times (adjusted intraclass correlation of 0.20, 95% CI: 0.12-0.29) was minor, and FPE rates showed no difference between-centers (adjusted intraclass correlation of 0.00, 95% CI: 0.00-0.01).

**Table 2:** Procedural Details and Outcomes.

| **Full Carrier Cohort** | **n = 211** |
| --- | --- |
| **Anesthesia** |  |
| MAC | 70 [33.2%] |
| GA | 141 [66.8%] |
| **Access** |  |
| Femoral | 189 [89.6%] |
| Radial | 22 [10.4%] |
| **Median Aspiration Catheter ID** | 71 [71-72] |
| **Aspiration Catheter ID** |  |
| 84-92" | 12 [5.7%] |
| 68-74" | 194 [91.9%] |
| <68" | 4 [1.9%] |
| **Carrier size** |  |
| Large | 35 [14.2%] |
| Medium | 174 [82.5%] |
| Small | 2 [1%] |
| **Microwire used** | 130 [61.6%] |
| 0.014" | 66 [50.8%] |
| 0.018" | 57 [43.8%] |
| 0.016" | 7 [5.4%] |
| **Microwire Not Used** | 81 [38.4%] |
| **Time to Clot with Wire** | 13 mins [9-19.75] |
| **Time to Clot Without Wire** | 12 mins [8-17] |
| **Combined Technique** | 21 [10%] |
| **Puncture to Clot** | 12 mins [9-19] |
| **Puncture to Reperfusion** | 20 mins [13-32] |
| **mFPE** | 157 [74.4%] |
| **FPE** | 107 [50.7%] |
| **Final TICl 2c-3** | 147 [69.7%] |
| **Hemorrhage** |  |
| Parenchymal Hematoma | 25 [11.8%] |
| Subarachnoid Hemorrhage | 14 [6.6%] |
| **Complications** |  |
| Dissection | 1 [0.4%] |
| Perforation | 1 [0.4%] |
| **Discharge mRS 0-2** | 66 [31.2%] |
| **90-day mRS 0-2** | 67 of 168 [39.8%] |
| **Mortality** | 35 of 168 [20.8%] |

Legend: TICI - Thrombolysis in Cerebral Infarction, FPE - First Pass Effect, mFPE - modified FPE, mRS - modified Rankin Scale.

Successful clot engagement was achieved in 202 cases (95.7%) using two carrier delivery methods. The microwire-free (“wireless”) delivery technique allowed clot access in 79/81 cases (39.1%), while microwire-assisted (“wired”) Carrier delivery in 123/130 cases (60.9%) - the rate of conversion from wireless to wired was not recorded.

### Safety and Clinical Outcomes

The rates of parenchymal hematomas and subarachnoid hemorrhage were 11.8% and 6.6%, respectively(**Table 2**). Complications included one arterial dissection in the extracranial vertebral with the use of wireless technique, and one perforation noticed after pull-back of a SR deployed via Carrier after wireless MCA M1 thrombus crossing. The rate of modified Rankin Scale 0-2 was 67/168 (39.8%) and mortality 35/168 (16.6%) at 90-days.

### Sensitivity Analysis: Impact of Cavernous Tortuosity

The baseline characteristics and imaging findings were comparable between patients with type 1-2 vs type 3-4 cavernous tortuosity (**Table 3**). The adjusted analysis indicated that the degree of tortuosity did not have a significant impact on the co-primary (β=2.75, t=1.51, p=0.07 for time-to-clot, and β=-0.17, SE=0.33, p=0.59 for FPE) and secondary endpoints (β=6.17, t=1.95, p=0.052 for time-to-reperfusion, β=0.37, SE=0.4, p=0.35 for mFPE, and β=0.05, SE=0.52, p=0.92 for PH).

**Table 3:** Comparison of performance in different degrees of carotid siphon tortuosity.

| **Carrier [n = 211]** | **Type 1-2 Siphon**<br>**n=143** | **Type 3-4 Siphon**<br>**n=59** | **p-value** |
| --- | --- | --- | --- |
| **Baseline Characteristics** |  |  |  |
| **Age [years]** | 70 [60-78] | 73 [63.5-81.5] | 0.22 |
| **Hypertension** | 110 [76.2%] | 44 [74.6%] | 0.80 |
| **Dyslipidemia** | 78 [54.5%] | 36 [61%] | 0.49 |
| **Diabetes** | 51 [35.7%] | 21 [35.6%] | 1.00 |
| **Atrial Fibrillation** | 52 [36.4%] | 20 [33.9%] | 0.80 |
| **Smoking [Current]** | 28 [19.6%] | 9 [15.3%] | 0.60 |
| **NIHSS** | 16 [11-20] | 17 [11-22] | 0.30 |
| **ASPECTS** | 8 [7-9] | 8 [7-10] | 0.40 |
| **IV Thrombolysis** | 38 [26.6%] | 15 [25.4%] | 1.00 |
| **Vessels** |  |  |  |
| Internal carotid artery-T | 20 [14%] | 6 [10.2%] | 1.00 |
| MCA M1 | 83 [58%] | 39 [66.1%] | 1.00 |
| MCA M2 | 12 [8.4%] | 8 [13.6%] | 1.00 |
| Basilar | 2 [1.4%] | 0 [0%] | 1.00 |
| **Procedural Details** |  |  |  |
| **Anesthesia** |  |  |  |
| MAC | 50 [35%] | 20 [33.9%] | 1.00 |
| GA | 93 [65%] | 39 [66.1%] | 1.00 |
| **Access** |  |  |  |
| Femoral | 134 [93.7%] | 49 [83%] | 0.07 |
| Radial | 9 [6.3%] | 10 [17%] |  |
| **Median Aspiration Catheter ID** | 71 [71-73] | 71 [70-72] | 0.80 |
| **Super Large Bore** | 9 [6.3%] | 3 [5.1%] | 0.90 |
| **Carrier size** |  |  |  |
| Large | 20 [14%] | 15 [25.4%] | 0.30 |
| Medium | 120 [83.9%] | 44 [74.6%] | 0.50 |
| Small | 2 [1.4%] | 0 [0%] | 1.00 |
| **Wireless Navigation** | 55 [38.5%] | 24 [40.7%] | 0.80 |
| **Puncture to Clot time** | 12 mins [8.25-17.75] | 13.5 mins [9.25-21.75] | 0.07 |
| **Puncture to reperfusion** | 19 mins [IQR] | 22 mins [14.5-31.5] | 0.05 |
| **mFPE** | 102 [71.3%] | 47 [79.7%] | 0.29 |
| **FPE** | 71 [49.6%] | 30 [50.8%] | 1.00 |
| **Final TIC1 2c-3** | 100 [69.9%] | 40 [67.8%] | 0.89 |
| **Imaging and Clinical Outcomes** |  |  |  |
| **Hemorrhage** |  |  |  |
| Parenchymal Hematoma | 18 [12.5%] | 7 [11.8%] | 1.00 |
| Subarachnoid Hemorrhage | 11 [7.7%] | 3 [5.1%] | 0.70 |
| **Discharge mRS 0-2** | 45 [31.5%] | 17 [28.8%] | 0.80 |
| **90-day mRS 0-2** | 44 of 111[39.6%] | 19 of 49 [38.7%] | 0.90 |
| **Mortality** | 20 of 111 [18%] | 15 of 49 [30.61%] | 0.08 |

Legend: NIHSS - National Institutes of Health Stroke Scale, ASPECTS – Alberta Stroke Program Early CT Score, MCA- Middle cerebral artery, TICI – Thrombolysis in Cerebral Infarction, FPE – First Pass Effect, mFPE – modified First Pass Effect, mRS – modified Rankin Scale

### Pre-specified Comparative Analysis-Carrier DAC vs Standard Microcatheter/Microwire

At the single center cohort, of 665 thrombectomies within the study period, 242 underwent thrombectomy for proximal occlusions using upfront contact aspiration (41 using carrier and 147 standard microcatheter technique) (**Table 4**). The demographics were comparable between groups. Stroke severity, ASPECTS, rates of IV thrombolysis and occlusion-site were similar. Carrier cases more commonly used radial access (14 vs 3%; p<0.01). The median aspiration catheter ID was 0.071”[0.070-0.071”] vs 0.071”[0.071-0.071”]; p<0.01 for the Carrier and microcatheter groups, respectively.

**Table 4:** Comparison of Carrier vs Microcatheter Delivery of Contact Aspiration.

|  | **Non-Carrier**<br>n=147 | **Carrier**<br>n=41 | **p-value** |
| --- | --- | --- | --- |
| **Age [years]** | 65 [55.5-74.5] | 69 [58-76] | 0.39 |
| **Hypertension** | 102 [69.4%] | 27 [65.9%] | 0.80 |
| **Dyslipidemia** | 30 [20.4%] | 8 [19.5%] | 1.00 |
| **Diabetes** | 35 [23.8%] | 9 [22%] | 0.90 |
| **Atrial Fibrillation** | 34 [23.1%] | 9 [22%] | 1.00 |
| **Smoking [Current]** | 30 [20.4%] | 8 [19.5%] | 0.90 |
| **NIHSS** | 17 [12-22] | 18 [13-22] | 0.23 |
| **ASPECTS** | 8 [7-9] | 8 [7-9] | 0.34 |
| **IV Thrombolysis** | 48 [32.6%] | 12 [29.3%] | 0.80 |
| **Occlusion Site** |  |  |  |
| Internal carotid artery-T | 36 [24.5%] | 6 [14.6%] | 0.24 |
| MCA M1 | 79 [53.7%] | 29 [70.7%] | 0.07 |
| MCA M2 | 20 [13.6%] | 3 [7.3%] | 0.40 |
| Basilar | 12 [8.1%] | 3 [7.3%] | 1.00 |
| **Access** |  |  | 0.80 |
| Femoral | 142 [96.6%] | 35 [85.4%] | 0.01 |
| Radial | 5 [3.4%] | 6 [14.6%] | 0.01 |
| **Median Aspiration Catheter ID** | 71 [71-71] | 71 [70-71] | <0.01 |
| **Super Large Bore Catheters** | 32 [21.8%] | 4 [9.8%] | 0.10 |
| **Time to clot** | 13 mins [9.75-17.25] | 8 mins [6-11] | <0.01 |
| **Time to Reperfusion** | 23 mins [16-36.75] | 15 mins [10-28] | 0.03 |
| **mFPE** | 89 [60.6%] | 33 [80.5%] | 0.009 |
| **FPE** | 60 [40.8%] | 23 [56.1%] | 0.10 |
| **Final TICl 2c-3** | 111 [75.5%] | 36 [87.8%] | 0.10 |
| **Hemorrhage** |  |  |  |
| Parenchymal Hematoma | 19 [12.9%] | 3 [7.3%] | 0.40 |
| Subarachnoid Hemorrhage | 11 [7.5%] | 4 [9.7%] | 0.80 |
| **Discharge mRS 0-2** | 36 [24.4%] | 16 [39%] | 0.10 |
| **90-day mRS 0-2** | 51 of 130 [39.2%] | 19 [46.3%] | 0.20 |
| **Mortality** | 29 of 130 [22.3%] | 3 [7.3%] | 0.10 |

Legend: NIHSS - National Institutes of Health Stroke Scale, ASPECTS – Alberta Stroke Program Early CT Score, MCA – Middle Cerebral Artery, TICl – Thrombolysis in Cerebral Infarction, FPE – First Pass Effect, mFPE – modified First Pass Effect, mRS – modified Rankin Scale

#### Primary endpoints

The puncture-to-clot engagement was faster for Carrier cases after adjustments (8 [6-11] vs 13 [9-17] minutes; β = −5.80, SE = 2.34, *t* = −2.47, *p*=0.01) while the Carrier FPE rate was numerically higher compared to non-Carrier cases (56.1% vs 40.8%; β = 0.84, SE = 0.43, *t* = 1.95, *p*=0.05).

#### Secondary endpoints

the adjusted time-to-reperfusion times were shorter for the Carrier compared to the non-Carrier group (15 [10-28] vs 23 [16-36] min; β = -9.8, SE = 4.42, t = -2.22, p=0.02) while the Carrier mFPE rate was higher compared to non-Carrier cases (80.5% vs 60.6%; β = 1.66, SE = 0.55, *t* = 2.96, *p*=0.003). No association between Carrier and the likelihood of parenchymal hematoma (β = -0.3, SE = 0.73, p=0.59) was noted. Other radiological and clinical outcomes are listed in **Table 4**.

## DISCUSSION

This multicenter study of 211-patient demonstrates that the Carrier DAC is a safe and effective platform for large-bore aspiration catheter navigation during MT. The Carrier facilitated rapid aspiration catheter delivery with favorable procedural efficiency and no device related safety concerns.

The Delivery Assist Catheters represent an evolving class of devices designed to address the inherent technical challenges of navigating large-bore aspiration catheters through tortuous cerebrovascular anatomy. Traditional aspiration catheter delivery methods - including microwire/microcatheter combinations, macrowires, or directly advancing (“snaking”) the aspiration catheter - often result in suboptimal trackability and navigation due to size mismatch between the aspiration catheter inner diameter (ID) and the adjunctive device outer diameter (OD), creating a “ledge effect” impeding smooth catheter delivery .^10-15^ While smaller coaxial aspiration catheters can mitigate this ledge and provide structural support, they may add procedural complexity and increase cost.

Several DAC platforms have emerged as to overcome these limitations including, the Tenzing system (Route 92 Medical, San Mateo, CA), which has been demonstrated in the SummitMAX randomized clinical trial (RCT) to allow for clot access with super-bore catheters in 89.2% of the cases. The SendIT (Penumbra, Alameda, CA), Fast Pass (Stryker, Fremont, CA), Innerglide(Cerenovus, Irvine, CA) and the Zipline (Perfuze, Galway, Ireland) were also developed to enhance deliverability of aspiration catheters in challenging anatomy.^18-21,23^ The Carrier DAC similarly addresses these navigational challenges through a design optimized for rapid large-bore catheter delivery while having the of preserving procedural adaptability, including potential compatibility with certain stent-retrievers, although not indicated by instructions for use (IFU).^15,22^ The present study underscores the broader advantages of the DAC concept itself: providing more controlled and trackable navigation, mitigating anatomical challenges that have traditionally prolonged procedure times and limited reperfusion success.

Expedited reperfusion is strongly associated with improved functional outcomes.^27-28^ Early studies demonstrated that contact aspiration offered procedural time advantages over stent retrievers, with faster reperfusion times reported in trials such as ASTER and COMPASS.^3-4^ However, recent trials including VECTOR showed comparable procedural times between combined technique (stent retriever plus aspiration) and contact aspiration.^23^ Contemporary registry data from the SVIN Registry showed median puncture-to-reperfusion times of 33 minutes [IQR 23-52].^8^ The recently published initial experience with the Carrier DAC demonstrated mean puncture-to-clot access of 18.1 minutes (SD 9.4) and puncture-to-reperfusion of 34.8 minutes (SD 25.7).^22^ The SUMMIT MAX trial evaluating the HiPoint super large bore system (0.088” ID) with Tenzing delivery-assist technology did not report time from puncture-to-clot or to-reperfusion for comaparison.^18^ In the present multicenter study of 211 Carrier-mediated thrombectomies, the median puncture-to-clot was 12 minutes (IQR 9-19) with median puncture-to-reperfusion of 20 minutes (IQR 13-32). In order to define the potential relative benefit of the DAC, a comparative analysis was performed including 242 patients undergoing upfront contact aspiration (41 Carrier versus 147 standard microcatheter technique), demonstrating that Carrier use significantly reduced puncture-to-clot engagement time after adjustments. These findings suggest that newer DACs could potentially reestablish contact aspiration’s procedural efficiency in current practice.

FPE represents a critical procedural endpoint strongly associated with improved clinical outcomes and reduced procedural complications.^25^ Larger aspiration catheter inner diameters have been associated with not only higher first-pass effect rates but also shorter reperfusion times, despite the larger outer diameter.^8,26^ Contemporary benchmarks demonstrate variable FPE rates with contact aspiration: the VECTOR clinical trial reported 30% FPE rates with aspiration alone, the SVIN Registry showed 34%, while the RESTORE post market study with the use of the Sofia 6F catheter reported 41.5% .^8,,26-27^ The SUMMIT MAX randomized clinical trial reported 46% FPE.^18^ Although DAC platforms are primarily designed to optimize catheter navigation and facilitate delivery of these larger-bore catheters, emerging evidence suggests they may also enhance reperfusion efficacy.^21.29^ The initial feasibility study reported 39.5% FPE, and our expanded multicenter cohort demonstrated 50.7% FPE with 74.4% mFPE. Notably, the median aspiration catheter inner diameter in the multicenter cohort was 0.071” (IQR 0.071-0.072”), with only 5.7% utilizing super large bore catheters (ID≥0.084”), suggesting that the Carrier DAC may enhance FPE rates even with standard large-bore catheters.^22^ The mechanism underlying this enhanced reperfusion efficacy may relate to the DAC removal technique: upon clot contact, swift withdrawal of the DAC may create a “piston effect” that promotes clot initial corking within the aspiration catheter lumen. Additionally, the Carrier platform may optimize the advancement of the catheter tip onto the clot (if maintained proximal to the occlusion) or improve the catheter’s angle of engagement with the clot (if slightly pushing into the clot is technically necessary for delivery), facilitating more effective clot integration. These observations suggest that while DAC technology fundamentally addresses the challenge of navigating tortuous anatomy to reliably deliver aspiration catheters to the target occlusion—particularly through difficult segments such as the ophthalmic ICA and carotid siphon—the platform may confer additional advantages in reperfusion efficacy through optimized clot engagement mechanics, without introducing safety concerns.

Overall, the Carrier-mediated thrombectomy demonstrated a favorable safety profile in this multicenter experience. Parenchymal hematoma was observed in 11.8% and subarachnoid hemorrhage in 6.6% in the present cohort. In the ASTER randomized trial, first-line contact aspiration was associated with parenchymal hematoma in 12.8% and SAH in 6.9%. Similarly, the recent VECTOR trial reported parenchymal hematoma rates of 13% and subarachnoid hemorrhage in 5% with aspiration first strategy.^3-5^ Taken together, the hemorrhagic profile observed in this study appears well aligned with controlled and contemporary data. Two complications occurred in the context of wireless carrier advancement, one being an extracranial vertebral dissection and the other an intracranial perforation (potentially related to the Carrier, to the stentretriever, or to reperfusion injury). Although uncommon, these observations highlight that advancement of the DACs without a guiding microwire may theoretically involve risk. While wireless lesion crossing may reduce procedural steps, careful case selection and operator judgment are warranted. Overall, DAC-related complications appear uncommon, as prior studies of the SendIT, Innerglide, Fastpass,Tenzig and Carrier reported no adverse events related to the device.^18-20,22-23,30^

This study has inherent limitations related to its retrospective design. The absence of core-lab imaging adjudication is another shortcoming. Catheter selection was operator-dependent, introducing confounding bias. The comparative analysis of Carrier and non-Carrier had a relatively small sample size and despite including identical catheter models on each group, did not control for the specific rate of usage of the different devices. Despite the rate of successful wireless technique navigation being significant, we cannot determine the rate of success since the conversion from wireless to wired was not recorded. Carrier-mediated thrombectomy in patients with more severe siphon tortuosity led to comparable rates of FPE as compared to more benign cavernous anatomy, while time from puncture-to-reperfusion was similar but nearly reached statistical significance (12 vs 13.5min; p=0.07). The use of Carrier for the delivery of SR is off-label and additional data would be required for potential validation.

The Carrier delivery-assist catheter facilitates efficient navigation of large-bore aspiration catheters in complex anatomy and is a promising strategy for reducing procedural time and preserving versatility for aspiration-first and combined technique strategies. Prospective studies are warranted.

## Data Availability

The data that support the findings of this study are available from the corresponding author upon reasonable request.

